# Impact of State Telehealth Parity Laws for Private Payers on Hypertension Management before and during the COVID-19 Pandemic

**DOI:** 10.1101/2023.11.16.23298658

**Authors:** Donglan Zhang, Jun Soo Lee, Adebola Popoola, Sarah Lee, Sandra L. Jackson, Lisa M. Pollack, Xiaobei Dong, Nicole L. Therrien, Feijun Luo

**Author notes:** Donglan Zhang and Jun Soo Lee are joint 1st authors with equal contributions. **Correspondence to:** Feijun Luo, PhD Economist Division for Health Disease and Stroke Prevention Centers for Disease Control and Prevention.

## Abstract

**BACKGROUND:** Telehealth has emerged as an effective tool for managing common chronic conditions such as hypertension, especially during the COVID-19 pandemic. However, the impact of state telehealth payment and coverage parity laws on hypertension management remains uncertain.

**METHODS:** Data from the Merative^TM^ MarketScan® Commercial Claims and Encounters Database from January 1, 2016 to December 31, 2021 were used to construct the study cohort. The sample included non-pregnant individuals aged 25–64 years with hypertension. We reviewed and coded telehealth parity laws related to hypertension management in all 50 states and the District of Columbia, distinguishing between payment parity laws and coverage parity laws. The primary outcomes were antihypertension medication use, measured by the average medication possession ratio (MPR), medication adherence (MPR ≥80%), and average number of days of drug supply. We used a generalized difference-in-difference (DID) design to examine the impact of these laws. Results were presented as marginal effects and 95% confidence intervals (CI).

**RESULTS:** Among 353,220 individuals, states with payment parity laws were significantly linked to increased average MPR by 0.43 percentage point (95% CI: 0.07 - 0.79), and an increase of 0.46 percentage point (95% CI: 0.06 - 0.92) in the probability of medication adherence. Payment parity laws also led to an average increase of 2.14 days (95% CI: 0.11 - 4.17) in antihypertensive drug supply, after controlling for state-fixed effects, year-fixed effects, individual sociodemographic characteristics and state time-varying covariates including unemployment rates, GDP per capita, and poverty rates. In contrast, coverage parity laws were associated with a 2.13-day increase (95% CI: 0.19 - 4.07) in days of drug supply, but did not significantly increase the average MPR or probability of medication adherence. In addition, telehealth payment or coverage parity laws were positively associated with the number of hypertension-related telehealth visits, but this effect did not reach statistical significance. These findings were consistent in sensitivity analyses.

**CONCLUSIONS:** State telehealth payment parity laws were significantly associated with greater medication adherence, whereas coverage parity laws were not. With the increasing adoption of telehealth parity laws across states, these findings may support policymakers in understanding potential implications on management of hypertension.

**Clinical Perspective:** *What Is New?:* Telehealth is an effective tool to manage hypertension and state-level telehealth parity laws can influence its application. Prior studies have not clearly differentiated between the impacts of payment parity and coverage parity. Using a quasi-experimental generalized difference-in-differences design, we assessed the effects of telehealth payment parity and coverage parity laws on hypertension management. Our study found that state telehealth payment parity laws were significantly associated with increased hypertension medication adherence, while coverage parity laws were not.

*What Are the Clinical Implications?:* The widespread adoption of telehealth payment parity laws may significantly impact hypertension management, during emergencies like the COVID-19 pandemic and beyond. Considering that hypertension impacts approximately half of the adult population, our study provides valuable insights into the potential benefits of telehealth parity laws for private payers in enhancing the management of hypertension. With the increasing adoption of telehealth parity laws across states, integrating telehealth into hypertension management holds significant implications for the evolving U.S. healthcare system in the digital age.

## INTRODUCTION

The COVID-19 pandemic brought about unprecedented changes in healthcare, leading to a rapid expansion of telehealth services to compensate for deferred medical care. Telehealth emerged as a critical and robust tool in managing prevalent chronic conditions like hypertension.^1, 2^ Telemedicine for hypertension is especially well-received, with one report claiming an average adherence to telemedicine-based hypertension management programs as high as 77%.^2^ However, telehealth adoption has been largely influenced by state policies, particularly those governing private payers’ (otherwise known as private insurance, primarily funded through benefits plans provided by employers) reimbursement. In contrast to public payers like Medicare and Medicaid, private payers covered over half of insured individuals.^3^ Despite the growing importance of telehealth, little is known about the impact of different state telehealth policies for private payers on hypertension management.

Telehealth parity requirements typically fall into two distinct types. The first type is “payment parity,” which requires that payments for telehealth services match those for in-person care. Payers must reimburse telehealth services at the same rate or amount as they would for in-person visits.^4, 5^ The second type is “coverage parity,” which mandates that services covered in-person must also be covered via telehealth, though not necessarily at an equal amount as in in-person care. ^4, 5^ This means the same services are eligible for coverage, but reimbursement rates may vary between telehealth and in-person visits. Previous research has explored the association of telehealth parity laws with various outcomes. For example, two studies found that utilization of telehealth services significantly increased in parity versus non-parity states.^6, 7^ In addition, studies have shown similar or better clinical outcomes for patients receiving telehealth care compared with in-person care in various clinical settings.^8, 9^ However, these studies often overlooked the differentiation between “payment parity” and “coverage parity” when investigating the effects of parity. Some states have coverage laws in place, indicating that telehealth services are eligible for coverage, but they might not mandate payment parity, and vice versa.^10^ The distinction between these types of parity is crucial, as it can significantly impact access to telehealth services and reimbursement rates for healthcare providers. Moreover, prior research on telehealth has been limited to other settings or services, such as mental health care and psychotherapy,^11, 12^ and has not specifically assessed hypertension treatment, particularly for private payers.

This study aims to fill a gap in the literature by separately investigating the impact of state telehealth payment parity laws and coverage parity laws on hypertension management before and during the COVID-19 pandemic. As outlined in Anderson’s behavioral model,^13^ insurance policy serves as a vital enabling factor for physicians to provide essential care to patients with chronic conditions and encourage patients to seek healthcare services. In the context of hypertension management, consistent follow-up for medication prescriptions, conducted via telehealth or in-person consultations, and adhering to antihypertensive medication regimens are key strategies to improve hypertension control.^14,15^ Thus, we hypothesize by providing insurance coverage for telehealth services, telehealth parity laws can enhance hypertension management by improving care and medication adherence. Our findings have important implications for policymakers, clinicians, and patients seeking to enhance the use of telehealth for hypertension management.

## METHODS

### Data Source

We used data from the Merative^TM^ MarketScan^®^ Commercial Claims and Encounters (CCAE) Database from January 1, 2016 to December 31, 2021.^16^ The MarketScan^®^ CCAE Database comprises patient-level administrative medical and outpatient drug claims, covering millions of U.S. individuals. The CCAE Database consists of more than 120 large employers and more than 30 health plans, with 27.5+ and 17.2+ million enrollees in 2016 and 2021, respectively. The CCAE Database includes claims data for individuals from all 50 U.S. states and the District of Columbia (DC), although the distribution is uneven across the regions. The inclusion of unique identifiers in the database allowed for continuous tracking of the same individuals over time. This study employed secondary data analysis, using de-identified information; thus, no review by the Institutional Review Board at New York University Grossman School of Medicine or the Centers for Disease Control and Prevention was deemed necessary. The authors cannot share the MarketScan® commercial claims database publicly due to the data user agreement, but the program codes will be available upon request to the corresponding author.

### Study Sample

The study sample included individuals aged 25–64 years as of December 31, 2021. To be included, individuals were required to have had a hypertension diagnosis (International Classification of Diseases, 10th Revision, Clinical Modification [ICD-10-CM] of I10-I15) during the years 2016–2017, identified as ≥1 inpatient or ≥2 outpatient visits at least 30 days apart (Appendix Table 1). Furthermore, individuals were required to have had ≥1 drug claim for an antihypertensive medication. We identified seven antihypertensive therapeutic classes, including ACE inhibitors, angiotensin receptor blockers, beta-blockers, calcium channel blockers, diuretics, renin-angiotensin system antagonists, and other antihypertensives, along with the corresponding generic drug names (Appendix Table 2).

**Table 1.**
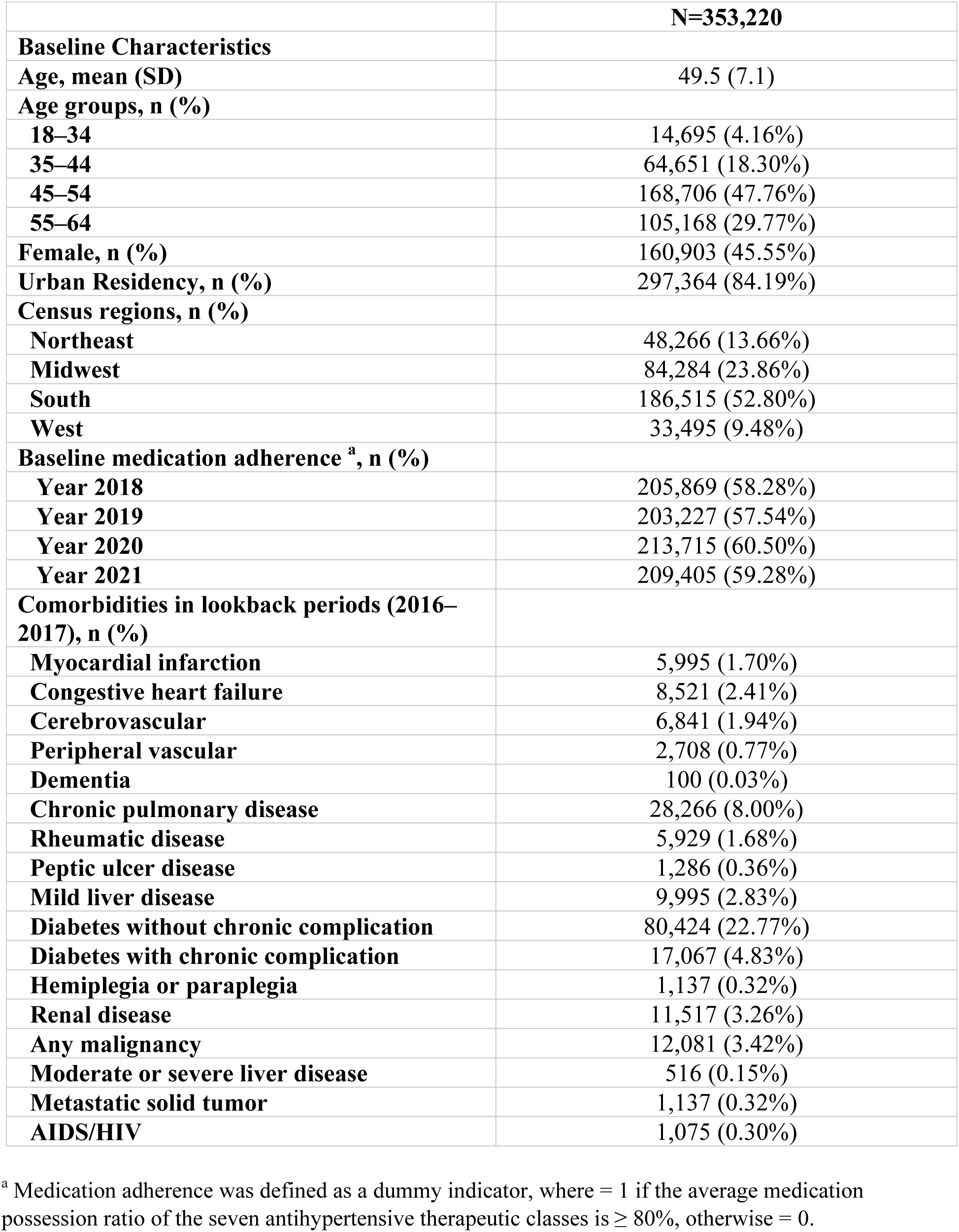
Baseline Characteristics of patients with hypertension in MarketScan® Commercial Claims and Encounters Database, 2016–2021.

|  | **N=353,220** |
| --- | --- |
| **Baseline Characteristics** |  |
| **Age, mean (SD)** | 49.5 (7.1) |
| **Age groups, n (%)** |  |
| **18–34** | 14,695 (4.16%) |
| **35–44** | 64,651 (18.30%) |
| **45–54** | 168,706 (47.76%) |
| **55–64** | 105,168 (29.77%) |
| **Female, n (%)** | 160,903 (45.55%) |
| **Urban Residency, n (%)** | 297,364 (84.19%) |
| **Census regions, n (%)** |  |
| **Northeast** | 48,266 (13.66%) |
| **Midwest** | 84,284 (23.86%) |
| **South** | 186,515 (52.80%) |
| **West** | 33,495 (9.48%) |
| **Baseline medication adherence <sup>a</sup>, n (%)** |  |
| **Year 2018** | 205,869 (58.28%) |
| **Year 2019** | 203,227 (57.54%) |
| **Year 2020** | 213,715 (60.50%) |
| **Year 2021** | 209,405 (59.28%) |
| **Comorbidities in lookback periods (2016–2017), n (%)** |  |
| **Myocardial infarction** | 5,995 (1.70%) |
| **Congestive heart failure** | 8,521 (2.41%) |
| **Cerebrovascular** | 6,841 (1.94%) |
| **Peripheral vascular** | 2,708 (0.77%) |
| **Dementia** | 100 (0.03%) |
| **Chronic pulmonary disease** | 28,266 (8.00%) |
| **Rheumatic disease** | 5,929 (1.68%) |
| **Peptic ulcer disease** | 1,286 (0.36%) |
| **Mild liver disease** | 9,995 (2.83%) |
| **Diabetes without chronic complication** | 80,424 (22.77%) |
| **Diabetes with chronic complication** | 17,067 (4.83%) |
| **Hemiplegia or paraplegia** | 1,137 (0.32%) |
| **Renal disease** | 11,517 (3.26%) |
| **Any malignancy** | 12,081 (3.42%) |
| **Moderate or severe liver disease** | 516 (0.15%) |
| **Metastatic solid tumor** | 1,137 (0.32%) |
| **AIDS/HIV** | 1,075 (0.30%) |

<sup>a</sup> Medication adherence was defined as a dummy indicator, where = 1 if the average medication possession ratio of the seven antihypertensive therapeutic classes is $\geq 80\%$ , otherwise = 0.

**Table 2.**
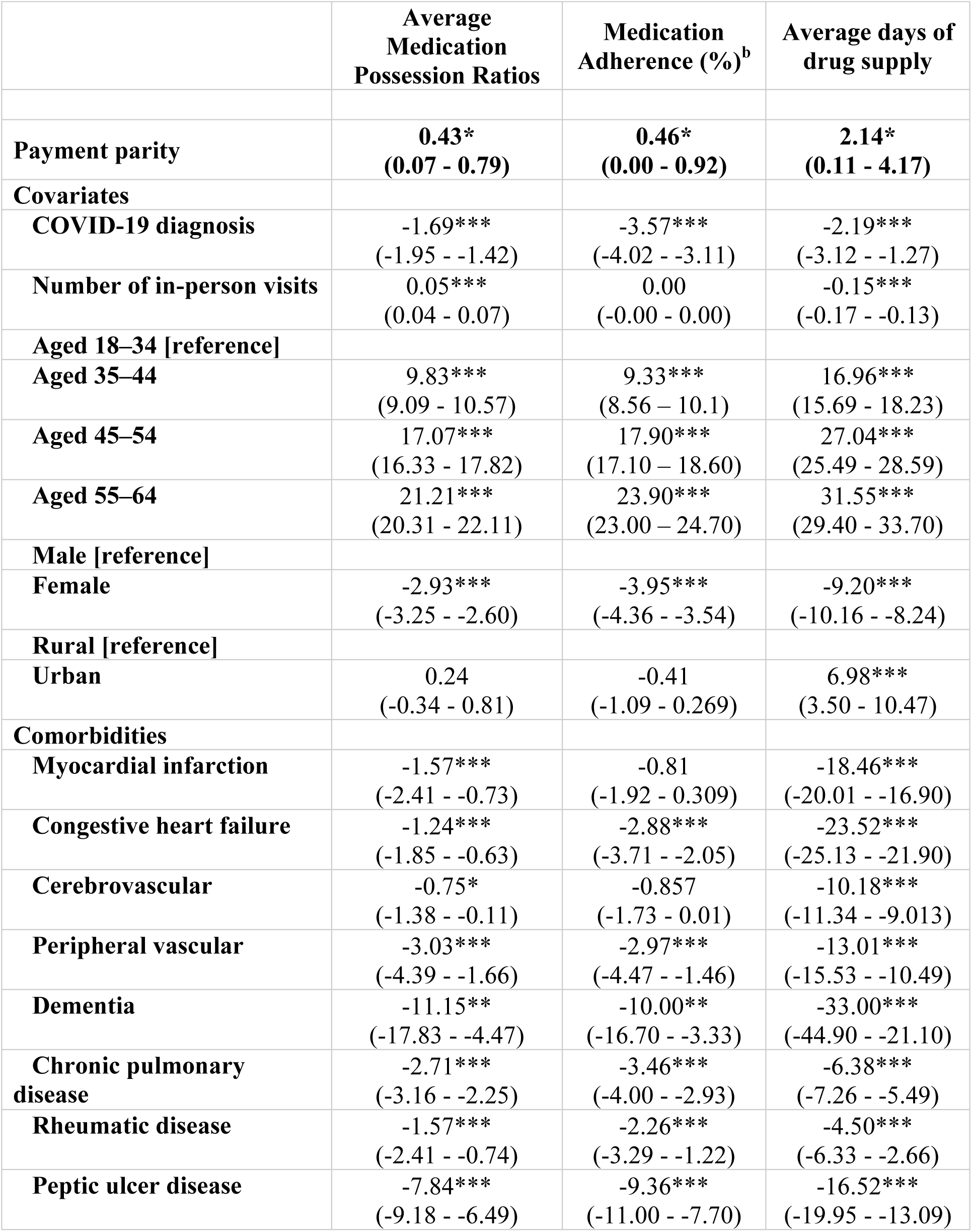
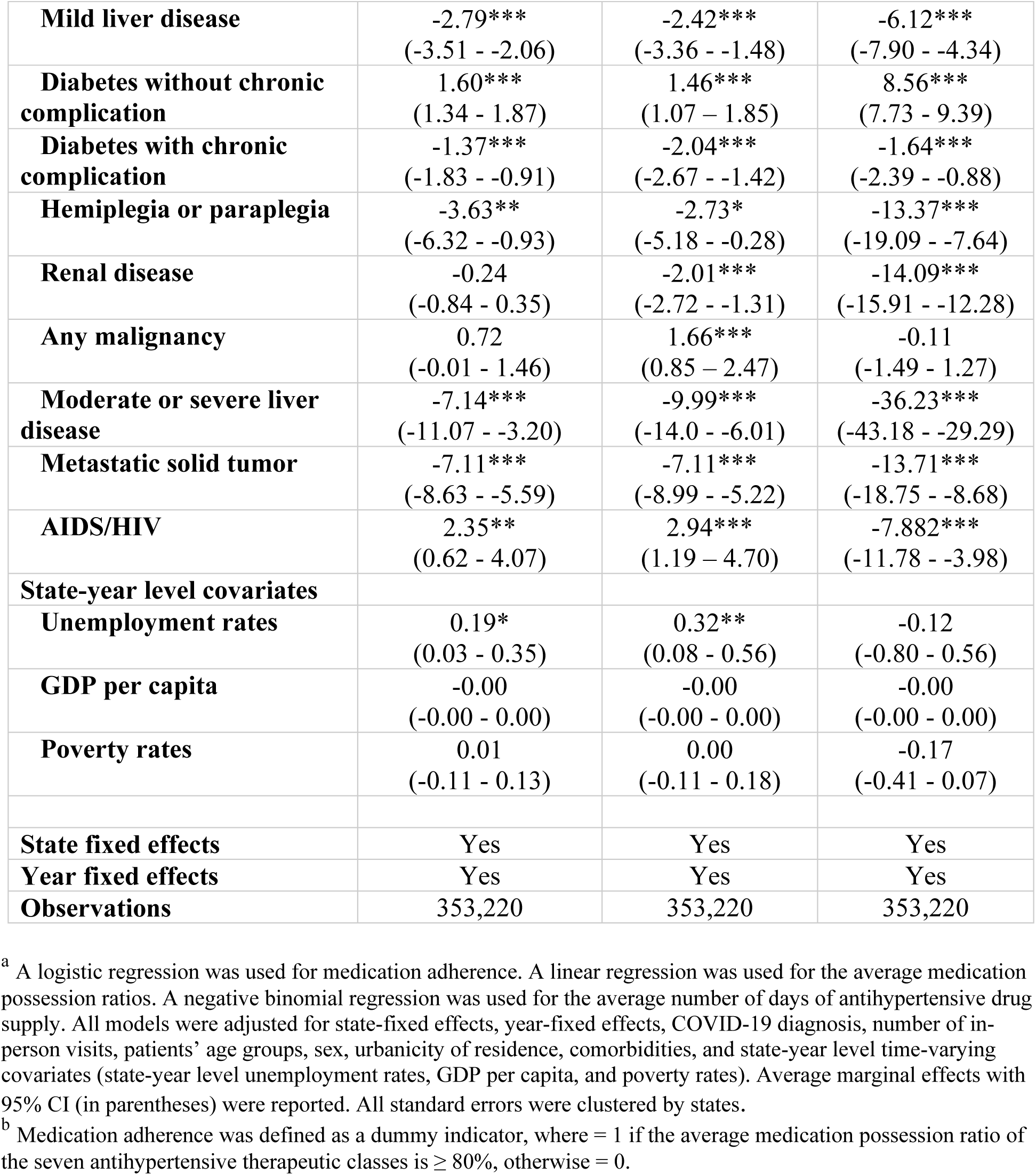
The association of <u>telehealth payment parity laws</u> with medication adherence, medication possession ratios, and average number of drug supply per antihypertensive drug, 2018–2021.^a^.

|  | **Average**<br>**Medication**<br>**Possession Ratios** | **Medication**<br>**Adherence (%)<sup>b</sup>** | **Average days of**<br>**drug supply** |
| --- | --- | --- | --- |
| **Payment parity** | **0.43\***<br>**(0.07 - 0.79)** | **0.46\***<br>**(0.00 - 0.92)** | **2.14\***<br>**(0.11 - 4.17)** |
| **Covariates** |  |  |  |
| **COVID-19 diagnosis** | -1.69***<br>(-1.95 - -1.42) | -3.57***<br>(-4.02 - -3.11) | -2.19***<br>(-3.12 - -1.27) |
| **Number of in-person visits** | 0.05***<br>(0.04 - 0.07) | 0.00<br>(-0.00 - 0.00) | -0.15***<br>(-0.17 - -0.13) |
| **Aged 18–34 [reference]** |  |  |  |
| **Aged 35–44** | 9.83***<br>(9.09 - 10.57) | 9.33***<br>(8.56 - 10.1) | 16.96***<br>(15.69 - 18.23) |
| **Aged 45–54** | 17.07***<br>(16.33 - 17.82) | 17.90***<br>(17.10 - 18.60) | 27.04***<br>(25.49 - 28.59) |
| **Aged 55–64** | 21.21***<br>(20.31 - 22.11) | 23.90***<br>(23.00 - 24.70) | 31.55***<br>(29.40 - 33.70) |
| **Male [reference]** |  |  |  |
| **Female** | -2.93***<br>(-3.25 - -2.60) | -3.95***<br>(-4.36 - -3.54) | -9.20***<br>(-10.16 - -8.24) |
| **Rural [reference]** |  |  |  |
| **Urban** | 0.24<br>(-0.34 - 0.81) | -0.41<br>(-1.09 - 0.269) | 6.98***<br>(3.50 - 10.47) |
| **Comorbidities** |  |  |  |
| **Myocardial infarction** | -1.57***<br>(-2.41 - -0.73) | -0.81<br>(-1.92 - 0.309) | -18.46***<br>(-20.01 - -16.90) |
| **Congestive heart failure** | -1.24***<br>(-1.85 - -0.63) | -2.88***<br>(-3.71 - -2.05) | -23.52***<br>(-25.13 - -21.90) |
| **Cerebrovascular** | -0.75*<br>(-1.38 - -0.11) | -0.857<br>(-1.73 - 0.01) | -10.18***<br>(-11.34 - -9.013) |
| **Peripheral vascular** | -3.03***<br>(-4.39 - -1.66) | -2.97***<br>(-4.47 - -1.46) | -13.01***<br>(-15.53 - -10.49) |
| **Dementia** | -11.15**<br>(-17.83 - -4.47) | -10.00**<br>(-16.70 - -3.33) | -33.00***<br>(-44.90 - -21.10) |
| **Chronic pulmonary**<br>**disease** | -2.71***<br>(-3.16 - -2.25) | -3.46***<br>(-4.00 - -2.93) | -6.38***<br>(-7.26 - -5.49) |
| **Rheumatic disease** | -1.57***<br>(-2.41 - -0.74) | -2.26***<br>(-3.29 - -1.22) | -4.50***<br>(-6.33 - -2.66) |
| **Peptic ulcer disease** | -7.84***<br>(-9.18 - -6.49) | -9.36***<br>(-11.00 - -7.70) | -16.52***<br>(-19.95 - -13.09) |
| **Mild liver disease** | -2.79***<br>(-3.51 - -2.06) | -2.42***<br>(-3.36 - -1.48) | -6.12***<br>(-7.90 - -4.34) |
| **Diabetes without chronic complication** | 1.60***<br>(1.34 - 1.87) | 1.46***<br>(1.07 - 1.85) | 8.56***<br>(7.73 - 9.39) |
| **Diabetes with chronic complication** | -1.37***<br>(-1.83 - -0.91) | -2.04***<br>(-2.67 - -1.42) | -1.64***<br>(-2.39 - -0.88) |
| **Hemiplegia or paraplegia** | -3.63**<br>(-6.32 - -0.93) | -2.73*<br>(-5.18 - -0.28) | -13.37***<br>(-19.09 - -7.64) |
| **Renal disease** | -0.24<br>(-0.84 - 0.35) | -2.01***<br>(-2.72 - -1.31) | -14.09***<br>(-15.91 - -12.28) |
| **Any malignancy** | 0.72<br>(-0.01 - 1.46) | 1.66***<br>(0.85 - 2.47) | -0.11<br>(-1.49 - 1.27) |
| **Moderate or severe liver disease** | -7.14***<br>(-11.07 - -3.20) | -9.99***<br>(-14.0 - -6.01) | -36.23***<br>(-43.18 - -29.29) |
| **Metastatic solid tumor** | -7.11***<br>(-8.63 - -5.59) | -7.11***<br>(-8.99 - -5.22) | -13.71***<br>(-18.75 - -8.68) |
| **AIDS/HIV** | 2.35**<br>(0.62 - 4.07) | 2.94***<br>(1.19 - 4.70) | -7.882***<br>(-11.78 - -3.98) |
| **State-year level covariates** |  |  |  |
| **Unemployment rates** | 0.19*<br>(0.03 - 0.35) | 0.32**<br>(0.08 - 0.56) | -0.12<br>(-0.80 - 0.56) |
| **GDP per capita** | -0.00<br>(-0.00 - 0.00) | -0.00<br>(-0.00 - 0.00) | -0.00<br>(-0.00 - 0.00) |
| **Poverty rates** | 0.01<br>(-0.11 - 0.13) | 0.00<br>(-0.11 - 0.18) | -0.17<br>(-0.41 - 0.07) |
| **State fixed effects** | Yes | Yes | Yes |
| **Year fixed effects** | Yes | Yes | Yes |
| **Observations** | 353,220 | 353,220 | 353,220 |

To ensure data quality and consistency, we excluded individuals who were not continuously enrolled in a health plan, had a pregnancy-associated diagnosis (Appendix Table 1),^17, 18^ or were covered by capitated health insurance (as healthcare utilization and costs are not fully captured among those with capitated health insurance) during our study period (2016 to 2021). Participants were also excluded if any state identifiers were missing or unknown.

### State Telehealth Parity Laws

Using publicly accessible sources (i.e., state agency websites and national organization websites, such as the Center for Connected Health Policy), we reviewed and collected telehealth parity laws (legislations, regulations, executive orders, and agency policies) in the 50 states and Washington, D.C. in effect from January 1, 2018, to December 31, 2021. To ensure that the study adequately assessed the impact of state telehealth parity laws relevant to preventing, treating, and managing hypertension for individuals enrolled in private insurance, we focused on laws that apply to the provision of primary care services and relevant specialty care. These include cardiology, neurology, medication management, cardiac rehabilitation, telestroke, and chronic disease management.

For each state, relevant laws were independently coded by two researchers, and discrepancies in coding were resolved through discussions and consensus. Coding of telehealth coverage and payment parity laws were based on the legal language in each state. A parity law was coded as “payment parity” if it expressly required private payers to reimburse telehealth services at the same rate as an in-person visit (e.g., reimbursement for services provided through telehealth must be equivalent to reimbursement for the same services provided in-person). Similarly, a parity law was coded as “coverage parity” if it contained language expressly requiring private payers to cover the same services for telehealth as if it would have been covered for an in-person visit (e.g., shall provide coverage for services provided via telehealth if it was covered in-person and shall be subject to the same terms and conditions, but coverage parity does not guarantee the same rate of payment).

The state law analysis did not assess laws for specialty services not commonly associated with hypertension, stroke, or cardiovascular disease. These include mental health services, hospice, and palliative care. The coded telehealth laws (Appendix Table 3) were then linked to our study sample using state identifiers.

**Table 3.** The association of <u>telehealth coverage parity laws</u> with medication adherence, medication possession ratios, and average number of drug supply per antihypertensive drug, 2018–2021.^a^.

|  | **Average**<br>**Medication**<br>**Possession Ratios**<br>**(%)** | **Medication**<br>**Adherence (%)<sup>b</sup>** | **Average days of**<br>**drug supply** |
| --- | --- | --- | --- |
| **Coverage parity** | 0.26<br>(-0.14 - 0.66) | 0.10<br>(-0.36 - 0.56) | **2.13\***<br>**(0.19 - 4.07)** |
| **Covariates** |  |  |  |
| **COVID-19 diagnosis** | -1.69***<br>(-1.96 - -1.42) | -3.57***<br>(-4.03 - -3.11) | -2.20***<br>(-3.13 - -1.27) |
| **Number of in-person visits** | 0.05***<br>(0.04 - 0.07) | -0.00<br>(-0.02 - 0.02) | -0.15***<br>(-0.17 - -0.13) |
| **Aged 18–34 [reference]** |  |  |  |
| **Aged 35–44** | 9.83***<br>(9.09 - 10.57) | 9.33***<br>(8.56 - 10.10) | 16.96***<br>(15.69 - 18.23) |
| **Aged 45–54** | 17.07***<br>(16.33 - 17.82) | 17.90***<br>(17.1 - 18.60) | 27.04***<br>(25.49 - 28.59) |
| **Aged 55–64** | 21.21***<br>(20.31 - 22.11) | 23.90***<br>(23.00 - 24.70) | 31.55***<br>(29.41 - 33.70) |
| **Male [reference]** |  |  |  |
| **Female** | -2.93***<br>(-3.25 - -2.60) | -3.95***<br>(-4.36 - -3.54) | -9.20***<br>(-10.16 - -8.24) |
| **Rural [reference]** |  |  |  |
| **Urban** | 0.24<br>(-0.34 - 0.81) | -0.41<br>(-1.09 - 0.27) | 6.98***<br>(3.50 - 10.46) |
| **Comorbidities** |  |  |  |
| **Myocardial infarction** | -1.57***<br>(-2.41 - -0.73) | -0.81<br>(-1.92 - 0.31) | -18.46***<br>(-20.01 - -16.90) |
| **Congestive heart failure** | -1.24***<br>(-1.85 - -0.63) | -2.88***<br>(-3.71 - -2.05) | -23.52***<br>(-25.13 - -21.90) |
| **Cerebrovascular** | -0.75*<br>(-1.38 - -0.11) | -0.86<br>(-1.73 - 0.01) | -10.18***<br>(-11.34 - -9.012) |
| **Peripheral vascular** | -3.03***<br>(-4.39 - -1.66) | -2.97***<br>(-4.47 - -1.46) | -13.01***<br>(-15.52 - -10.49) |
| **Dementia** | -11.15**<br>(-17.83 - -4.47) | -10.00**<br>(-16.70 - -3.33) | -33.01***<br>(-44.90 - -21.11) |
| **Chronic pulmonary disease** | -2.71***<br>(-3.16 - -2.25) | -3.46***<br>(-4.00 - -2.93) | -6.37***<br>(-7.26 - -5.49) |
| **Rheumatic disease** | -1.57***<br>(-2.41 - -0.74) | -2.26***<br>(-3.29 - -1.22) | -4.50***<br>(-6.33 - -2.66) |
| **Peptic ulcer disease** | -7.84***<br>(-9.18 - -6.49) | -9.36***<br>(-11.0 - -7.70) | -16.52***<br>(-19.95 - -13.08) |
| **Mild liver disease** | -2.79***<br>(-3.51 - -2.06) | -2.42***<br>(-3.36 - -1.48) | -6.12***<br>(-7.90 - -4.34) |
| **Diabetes without chronic complication** | 1.60***<br>(1.34 - 1.87) | 1.46***<br>(1.07 - 1.85) | 8.56***<br>(7.73 - 9.39) |
| **Diabetes with chronic complication** | -1.37***<br>(-1.83 - -0.91) | -2.04***<br>(-2.67 - -1.42) | -1.63***<br>(-2.39 - -0.88) |
| **Hemiplegia or paraplegia** | -3.63**<br>(-6.32 - -0.93) | -2.73*<br>(-5.18 - -0.28) | -13.36***<br>(-19.09 - -7.64) |
| **Renal disease** | -0.24<br>(-0.84 - 0.35) | -2.01***<br>(-2.72 - -1.31) | -14.09***<br>(-15.91 - -12.28) |
| **Any malignancy** | 0.72<br>(-0.01 - 1.46) | 1.66***<br>(0.846 - 2.47) | -0.11<br>(-1.49 - 1.27) |
| **Moderate or severe liver disease** | -7.14***<br>(-11.07 - -3.20) | -9.99***<br>(-14.00 - -6.01) | -36.23***<br>(-43.18 - -29.29) |
| **Metastatic solid tumor** | -7.11***<br>(-8.63 - -5.59) | -7.11***<br>(-8.99 - -5.22) | -13.71***<br>(-18.75 - -8.68) |
| **AIDS/HIV** | 2.35**<br>(0.62 - 4.07) | 2.94***<br>(1.19 - 4.70) | -7.88***<br>(-11.78 - -3.982) |
| **State-year level covariates** |  |  |  |
| **Unemployment rates** | 0.19*<br>(0.03 - 0.36) | 0.30*<br>(0.05 - 0.55) | -0.05<br>(-0.71 - 0.61) |
| **GDP per capita** | -0.00<br>(-0.00 - 0.00) | -0.00<br>(-0.00 - 0.00) | -0.00<br>(-0.00 - 0.00) |
| **Poverty rates** | -0.00<br>(-0.11 - 0.11) | 0.03<br>(-0.12 - 0.18) | -0.26*<br>(-0.49 - -0.03) |
| **State fixed effects** | Yes | Yes | Yes |
| **Year fixed effects** | Yes | Yes | Yes |
| **Observations** | 353,220 | 353,220 | 353,220 |

### Outcomes

The primary outcomes of the study included antihypertensive medication use, measured by the three indicators: 1) average medication possession ratio (MPR) of antihypertensive drugs, 2) medication adherence to antihypertensive drugs (defined as MPR ≥80%), and 3) average number of days of antihypertensive drug supply. Additionally, as secondary outcome measures, we included the number of hypertension-related and cardiovascular disease (CVD)-related telehealth visits.

To calculate the average MPR, we estimated the means of the MPRs for the seven antihypertensive therapeutic classes (Appendix Table 2). Each MPR was computed as the ratio of the total days of antihypertensive drug supply to the number of days in each year.^19^ A patient was assessed as adherent if the average MPR was ≥80%.^19^ The average days of drug supply referred to the average number of pills of medication supplied per antihypertensive prescription.

Telehealth outpatient visits were defined if outpatient visits were telehealth-related using place of service and procedure modifiers (Appendix Table 4). In-person outpatient visits were defined as non-telehealth-related outpatient visits. Telehealth visits can be complementary to in-person care whereby in-person visits might increase follow-up telehealth appointments, or telehealth visits might result in a necessary in-person visit.^20^ In addition, telehealth can be a substitute to in-person services, such as a telehealth visit substituting an in-person visit during the pandemic.^21^ Therefore, we adjusted for the number of in-person visits when assessing telehealth visits as an outcome, and vice versa. Hypertension-and CVD-related telehealth outpatient visits were identified if telehealth outpatient visits included the respective diagnosis (Appendix Table 1). All outcomes were assessed on a per patient per year basis, providing a comprehensive analysis of hypertension management and telehealth utilization.

### Statistical Analysis

To estimate the association between telehealth parity laws and hypertension management, we used a generalized difference-in-difference (DID) design with different treatment timing.^22^ Given that telehealth parity laws were in effect in different states during various time periods, we utilized the different timing of these laws to estimate the generalized DID effects. Within the generalized DID methodology, we used linear regression for average MPR, logistic regression for medication adherence, and negative binomial regression for numbers of days of antihypertensive drug supply; we used exponential hurdle^23^ model for number of hypertension-and CVD-related telehealth outpatient visits, since some patients may have zero telehealth visits in a year (e.g., excess zeros in the outcome measure).

In sensitivity analyses to test the influence of different model setups on the estimated results we used Probit and linear models for medication adherence. For the number of days of antihypertensive drug supply, we used a zero-inflated negative binomial model. This approach accommodates patients with zero days of drug supply by combining a logit model for zero outcomes and a negative binomial model for the average number of days of drug supply.

All models were adjusted for state-fixed effects, year-fixed effects, COVID-19 diagnosis (claims with ICD-10-CM of U07.1), number of in-person visits, age groups (18–34 [reference], 35–44, 45–54, and 55–64 years), sex (male [reference], female), urbanicity of residence (urban [reference], rural, per US Census Metropolitan Statistical Area [MSA] designation),^16^ and 17 Quan-Charlson comorbidities.^24^ Comorbidities were identified as present if individuals had ≥1 inpatient admission or ≥2 outpatient encounters (≥30 days apart) with the corresponding Quan-Charlson ICD-10-CM diagnoses. In addition, state-year level time-varying covariates such as state-year level unemployment rates, GDP per capita, and poverty rates were included in the models. We reported the average marginal effects along with 95% confidence intervals (CI). To account for potential clustering effects, all standard errors were clustered by state. All outcomes are measured on a per patient per year basis, except for the number of telehealth visits, measured as per 1,000 patients per year.

Two-tailed *p*-values < 0.05 were used to indicate statistical significance. All the analyses were conducted using Stata SE 17.0 statistical software (StataCorp, College Station, TX) and SAS version 9.4 (SAS 9.4, Cary, NC. SAS Institute Inc.).

## RESULTS

There were 353,220 included individuals (Figure 1). The sociodemographic characteristics of the study cohort at baseline are summarized in Table 1. The average age of patients with hypertension was 49.5 (SD, 7.1) years. Women comprised 45.55% of the cohort, and 84.19% lived in urban areas. Baseline medication adherence fluctuated between 57.54% and 60.50% from 2018 to 2021. The most prevalent comorbidities during the lookback period (2016– 2017) included diabetes without chronic complications (22.77%), chronic pulmonary disease (8.00%), diabetes with chronic complications (4.83%), any malignancy (3.42%), and renal disease (3.26%). The distribution of the patient cohort in each state is presented in Appendix Table 5.

**Figure 1.**
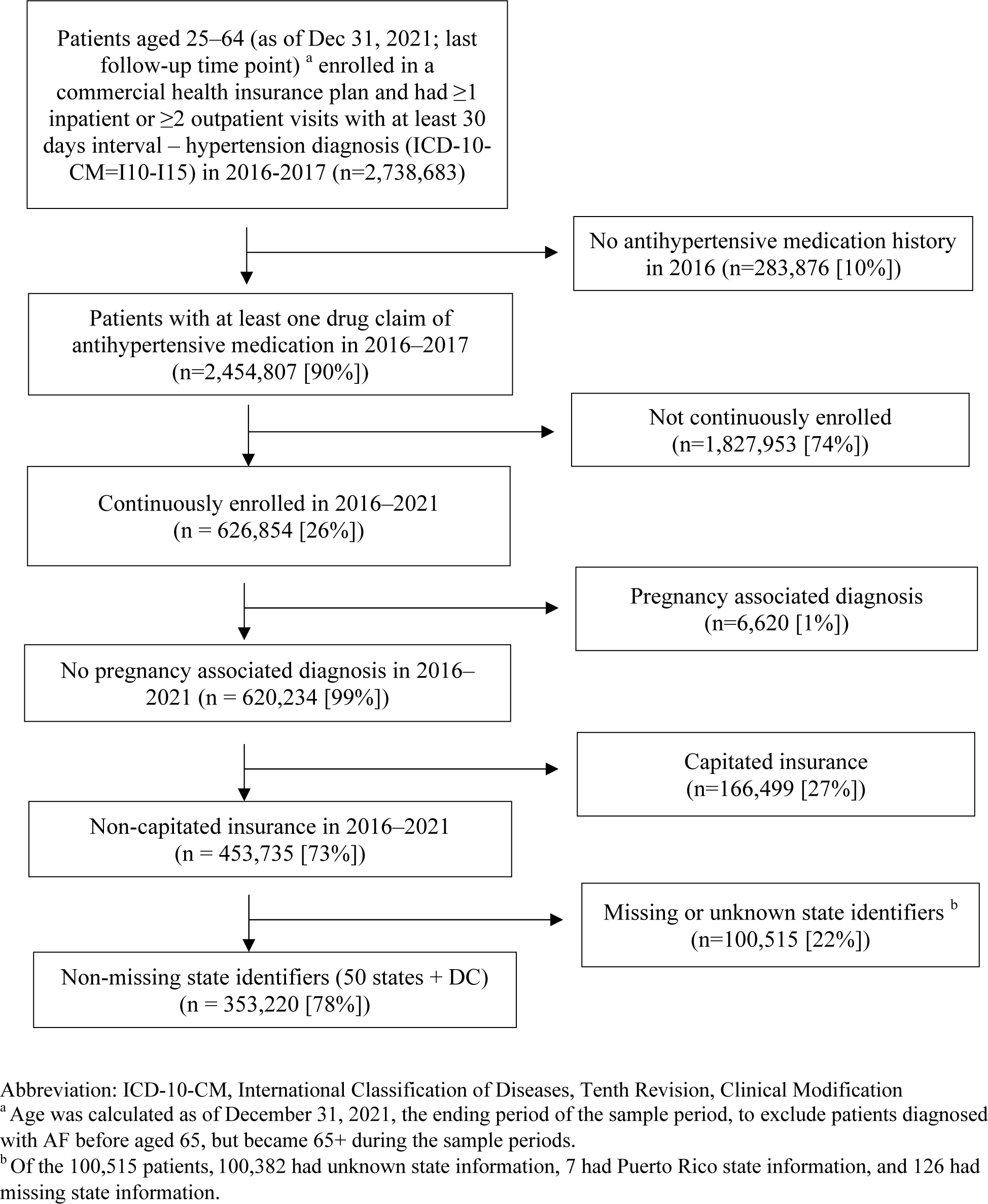
Study sample selection of patients with hypertension, MarketScan® Commercial Claims and Encounters Database, 2016–2021.

In 2018, one state had a payment parity law, 20 had coverage parity laws, and 6 had both. By 2021, these values had increased to 5 states with payment parity laws, 24 with coverage parity laws, and 16 with both.

After controlling for potential confounders, states with a payment parity law saw a statistically significant 0.43 percentage point (95% CI: 0.07 - 0.79) increase in average MPR among individuals with hypertension (Table 2). Furthermore, the payment parity law was significantly associated with a 0.46 percentage point (95% CI: 0.00 - 0.92) increase in the probability of medication adherence and a significant increase in average days of antihypertensive drug supply by 2.14 days (95% CI: 0.11 - 4.17) per patient per antihypertensive prescription.

Coverage parity laws were not significantly associated with average MPR or medication adherence, but were associated with a significant increase in average days of antihypertensive drug supply by 2.13 days (95% CI: 0.19 - 4.07) per patient per antihypertensive prescription.

Payment parity laws were associated with a significant increase in the number of hypertension-related telehealth visits by 2.61 visits per 1,000 patients (95% CI: 0.99 - 4.23) and CVD-related telehealth visits by 0.92 visits per 1,000 patients (95% CI: 0.23 - 1.61) (Appendix Table 6). On the other hand, coverage parity laws did not show a statistically significant association with the number of telehealth visits (Appendix Table 7).

In sensitivity analyses, we tested different model setups (Probit and linear regression) for the medication adherence measure, and the results remained largely unchanged (Appendix Table 8). Moreover, when using the zero-inflated negative binomial model for the average days of drug supply, the association with state payment parity was positive, but not statistically significant (Appendix Table 9).

## DISCUSSION

This study comprehensively assessed the potential impact of telehealth parity laws on hypertension management before and during the COVID-19 pandemic among private payers across the 50 states and DC. We distinguished between payment parity and coverage parity, as these different types of parity laws could have differential impact on patient access and behavior. Interestingly, the results revealed that state payment parity laws for hypertension-related telehealth services were significantly associated with increased average MPR, adherence to antihypertensive medications, and average days of drug supply. However, coverage parity laws were not found to be significantly associated with average MPR or medication adherence, but significantly associated with increased average days of drug supply. Furthermore, both types of parity laws did not significantly influence the numbers of hypertension-related telehealth visits or in-person visits during the study period. These findings remained robust after using alternative models in sensitivity analyses.

Applying a quasi-experimental generalized DID design, one key finding of our study was that telehealth payment parity laws were associated with significant improvements in medication adherence among patients with hypertension. It is possible that telehealth visits facilitated hypertension management by improving continuity of care, such as by alleviating patient concerns about exposure to SARS-CoV-2, the virus that causes COVID-19, during in-person office visits. In a recent study, Patel et al. (2020) estimated that the overall volume of care for hypertension dropped by 23.0% at the outset of the pandemic.^25^ Delayed care could lead to worsened outcomes for patients with poorly controlled hypertension, including heart attacks, coronary heart disease, strokes, and kidney damage.^26^ Consequently, telehealth use surged to ensure patient access to medical care without compromising patient and clinician safety. Though the marginal effect of 0.46 percentage point increase in the probability of medication adherence we observed seemed modest, considering that approximately 48 million individuals aged 20–64 have high blood pressure and are currently taking an antihypertensive medication, of whom 66.0% are covered by private insurance (as per NHANES 2017–2018 data),^27^ nationwide adoption of telehealth payment parity laws across all states could potentially result in approximately 145,728 more individuals achieving adherence to antihypertensive medications (MPR ≥ 80%). State telehealth payment parity laws were associated with immediate and positive impacts on populations affected by hypertension during the COVID-19 pandemic, especially given that telehealth usage has been associated with significant improvements in blood pressure control.^28^

Previous evidence-based studies suggest that an effective telehealth model for hypertension management should incorporate self-measured blood pressure monitoring or remote patient monitoring (RPM) that requires patients to record blood pressure outside of clinical settings and share the data with their clinicians.^29^ Another essential component of a telehealth visit includes consultations on the proper use of antihypertensive medications and adherence to medication regimens, and health education regarding lifestyle changes, often delivered through Internet video teleconferences on mobile devices.^2^ This approach may be combined with team-based care to provide this array of follow up services by a multidisciplinary team including physicians, nurses, health coaches, and pharmacists.^30^ But this combined approach may not always be practical, due to challenges such as patient access to digital platforms, reliable internet connection, and availability of compatible devices for vital sign measurement and transmission.^31–33^ To address these limitations, more simplified versions of telehealth, such as audio-only phone-based services, text messaging (SMS), or emails have also emerged as alternatives for communication that may still support treatment adherence and symptom surveillance.^34–36^ Some states have passed laws allowing reimbursement for audio-only services, primarily for the state Medicaid program, catering to the needs of patients with lower incomes who might lack smartphones or digital connectivity.^32,34^ Future studies could explore the influence of these laws on hypertension management.

We found that telehealth coverage parity laws were significantly associated with an increase in the average days of antihypertensive drug supply, but did not improve medication adherence. This could, in part, be attributed to telehealth coverage parity solely confirming insurance coverage without ensuring consistent reimbursement structures. Telehealth coverage parity alone can result in patients incurring higher out-of-pocket costs for telehealth services.

Another possibility is that coverage parity laws may influence clinician behavior. For instance, if the reimbursement rates for telehealth are lower, healthcare systems or clinicians may reduce the number of telehealth appointments offered and extend the duration of prescriptions for patients.

As shown in our study, more states adopted telehealth coverage parity or both parity laws rather than only payment parity laws. A recent review provides a comparative analysis of state-by-state adoption of telehealth parity, highlighting the need for alternative payment models, assessing resource utilization, and considering costs linked to virtual care.^8^ Certainly, looking beyond the pandemic, integrating telehealth into hypertension management without increasing utilization of telehealth may demand innovative payment models and care quality metrics.^37^

In the present study, we found a significant association between telehealth payment parity laws and the number of hypertension-and CVD-related telehealth visits. This finding was consistent with previous research. For example, a study analyzing 2010–2015 MarketScan® commercial claims data observed significant increases in outpatient telehealth visits in states with parity laws.^6^ Another study demonstrated enrollees in states with private payer telehealth laws were more likely to receive video assessments.^7^ However, our study did not find that telehealth coverage laws alone were significantly associated with the number of telehealth visits. Compared with prior studies, our study specifically examined the impact of these laws during the pandemic period, and focused on the laws relevant to hypertension and CVD management. These methodological and contextual differences may account for the differences in our findings.^25^

There are several key strengths of this study. First, unlike prior research that did not distinguish between the effects of payment parity laws and coverage parity laws, we contributed to the literature by evaluating the net impact of these two types of laws individually and teasing out their effects separately while providing a more comprehensive understanding of the impact of state laws governing private payer reimbursements for telehealth services. Second, while previous parity law studies predominantly centered around mental health services, our focus was on relevant laws affecting hypertension management and control, filling a critical research gap. Third, the variation in state policies across different years posed a challenge to the traditional DID analysis; thus, we used a generalized DID approach, which is a more advanced and flexible method to evaluate policy impacts.

There are a few limitations in our study. First, for the state law analysis, we identified and collected telehealth parity laws from secondary sources. Some relevant laws may have been missed because they were unavailable, used vague language, or were missing key information (such as legal citations). In instances where effective dates were missing, the dates were represented by a proxy date using January 1 of the respective year. Nevertheless, we believe that this should not significantly affect our analysis. For instance, if the law was implemented at the end of the year, our analysis was based on the data for that specific year, which may even make our estimates conservative. Second, some states might have implemented temporary telehealth COVID-19 emergency policies that were difficult to search for online, or may have expired during the study period. However, we did not include these policies in the treatment group and focused our analysis on permanent laws rather than temporary policies. Third, apart from telehealth parity laws, another important regulatory aspect at the state level is licensure requirements, ^38^ which can potentially influence the impact of parity laws on hypertension management. Some states have adopted interstate telemedicine laws, while others mandate registration for such practices.^3,8,9^ Language barriers and costs of telehealth infrastructure may hinder implementation. Additionally, varying state policies impact the rates of hospital telehealth adoption,^3^ and private payer reimbursement policies correlating with higher adoption rates. None of these factors were considered in the current analysis, as they fell outside the scope of the study. Fourth, although the MarketScan® commercial claims database provides a national sample that has comprehensive geographical representation covering all 50 states and D.C., there might be variations in the study population selected from different states. Nevertheless, this limitation has been mitigated through the adjustment for state fixed effects and state-level time-varying variables. Fifth, we did not assess whether participants relocated from one state to another during the study period. Since our sample selection criteria required continuous enrollment of participants during the study period, participants who moved were likely excluded from our study cohort.

## CONCLUSION

Our findings suggest that state telehealth payment parity laws may be significantly associated with hypertension treatment based on increased related telehealth visits and medication adherence. However, coverage parity laws alone do not have a significant association with telehealth services or medication adherence. Future studies could assess the associations of state telehealth laws for public payers and uninsured groups, as well as how licensure requirements are associated with hypertension management. Given that hypertension affects roughly half of the adult population, our study holds significant implications for the evolving U.S. healthcare system in the digital age.^39^ Our results offer insights into the potential role of telehealth parity laws for private payers in facilitating hypertension management.

## Disclaimer

The findings and conclusions in this report are those of the authors and do not necessarily represent the official position of the Centers for Disease Control and Prevention. Use of trade names and commercial sources is for identification only and does not imply endorsement by the U.S. Department of Health and Human Services.

## Data Availability

The authors cannot share the MarketScan° commercial claims database publicly due to the data user agreement, but the program codes will be available upon request to the corresponding author.

## Acknowledgments

The authors are genuinely grateful to Michael Schooley (Centers for Disease Control and Prevention) for his guidance, suggestions, and article review. The authors also thank researchers at NORC at the University of Chicago who assisted with collection and coding of state telehealth laws.

---

**Appendix Table 1.** ICD-10-CM diagnosis codes for hypertension and ICD-10-CM diagnosis, procedure, and DRG codes for pregnancy.

|  | **ICD-10-CM** | **DRG** | **ICD-10-PCS** |
| --- | --- | --- | --- |
| **Hypertension** | I10-I15 | NA | NA |
| **Cardiovascular disease** | I00-I78 | NA | NA |
| **Pregnancy** | O00-O99, O9A1-O9A5, Z33, Z34, Z36, Z37, Z3201, Z322, Z39, F53, A34 | 765, 766, 767, 768, 769, 770, 771, 772, 773, 775, 776, 777, 779, 780, 781, 782 | 10A0, 10D00Z0, 10D00Z1, 10D00Z2, 10D07Z3, 10D07Z4, 10D07Z5, 10D07Z6, 10D07Z7, 10D07Z8, 10E0XZZ |

Abbreviations: DRG, diagnosis-related group; ICD-10-CM, International Classification of Diseases, Tenth Revision, Clinical Modification; ICD-10-PCS, International Classification of Diseases, Tenth Revision, Procedure Coding System
NA indicates not available.

**Appendix Table 2.**
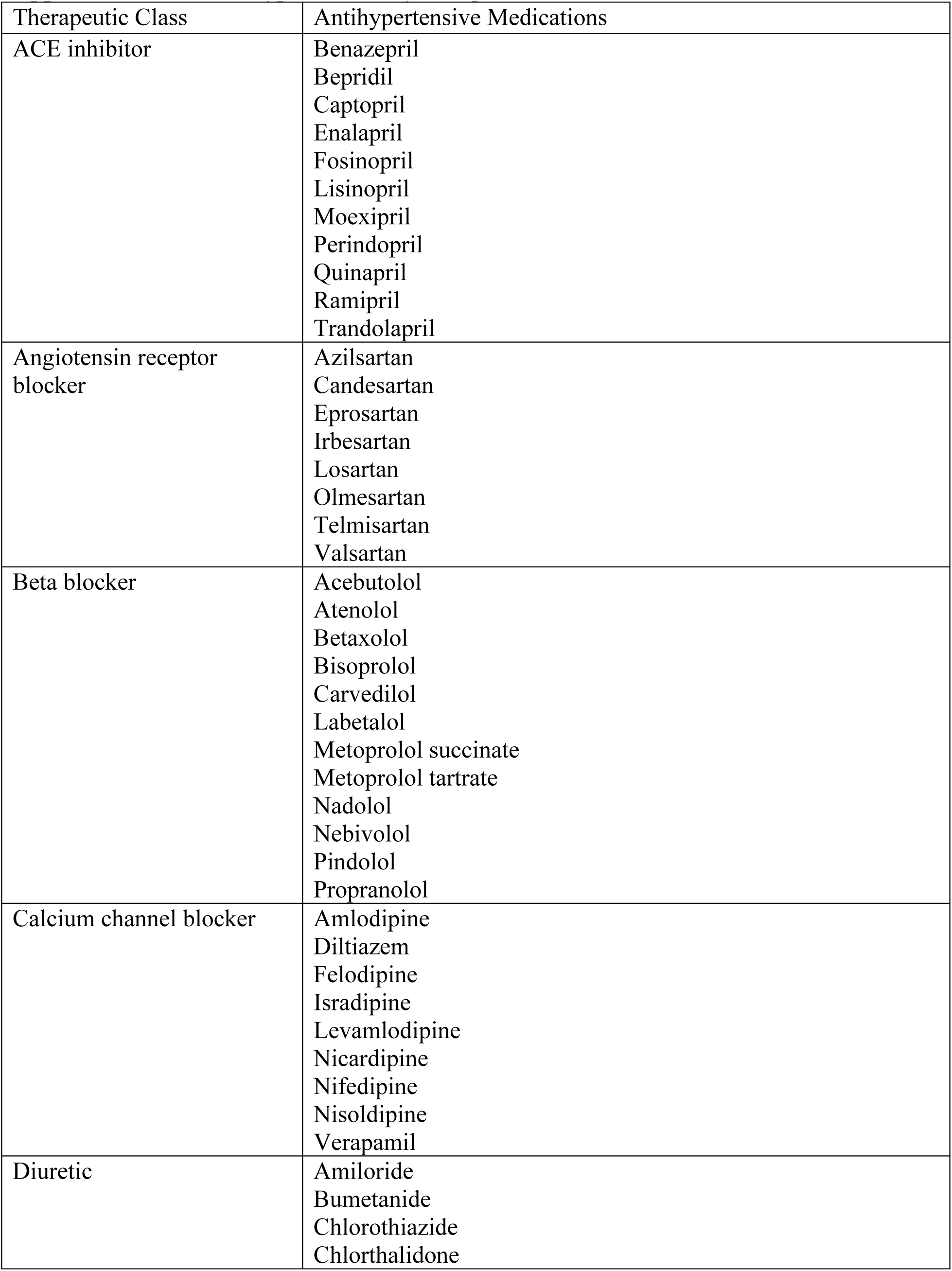
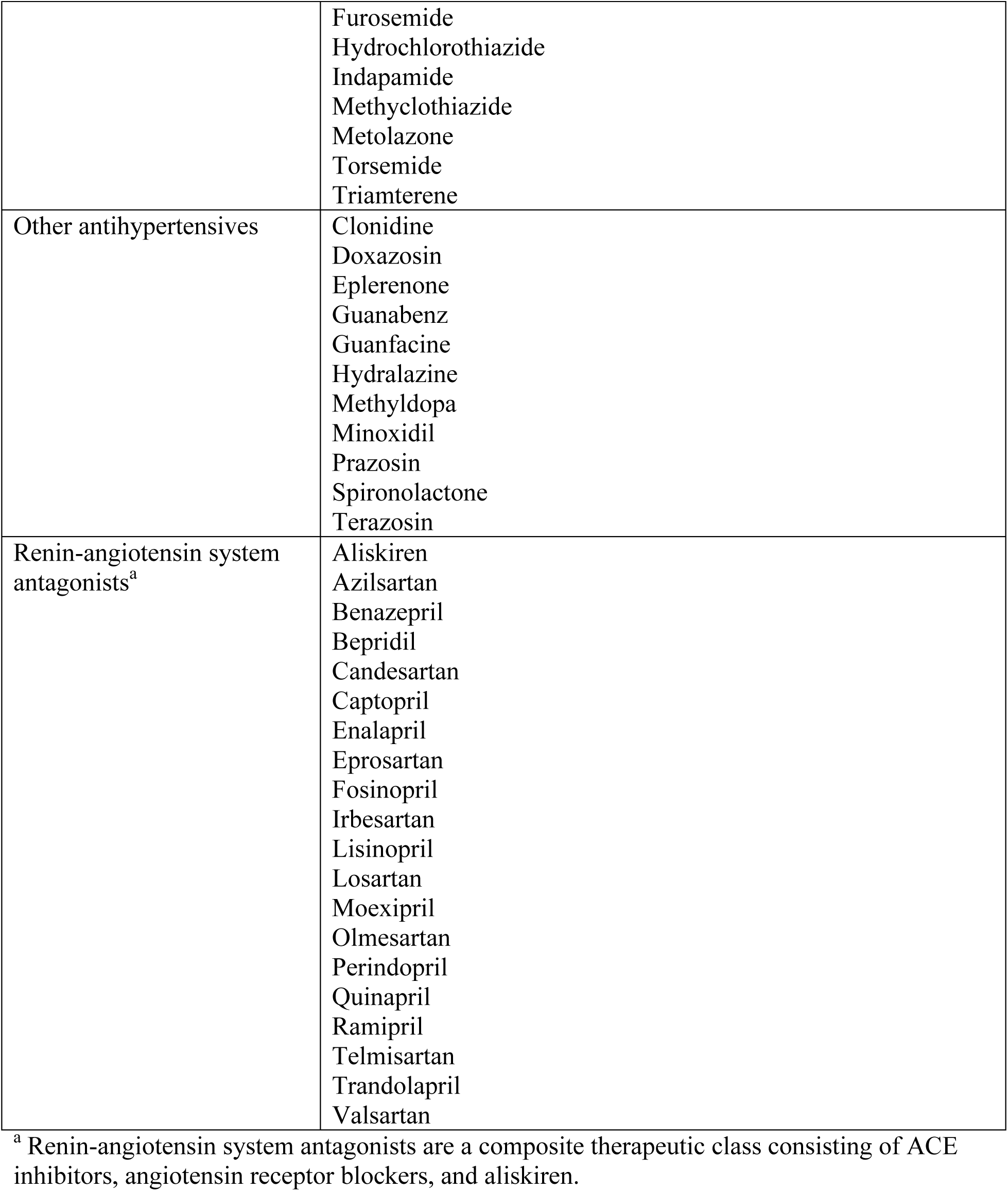
Antihypertensives by therapeutic class.

**Appendix Table 3.** State telehealth payment and coverage parity laws in effect January 1, 2018 – December 31, 2021.

| **State** | **2018** | **2019** | **2020** | **2021** |
| --- | --- | --- | --- | --- |
| **Alabama** | None | None | None | None |
| **Alaska** | None | None | Coverage | Coverage |
| **Arizona** | Coverage | Coverage | Coverage | Both |
| **Arkansas** | Both | Both | Both | Both |
| **California** | None | None | None | Both |
| **Colorado** | Coverage | Coverage | Both | Both |
| **Connecticut** | Coverage | Coverage | Coverage | Both |
| **Delaware** | Both | Payment | Both | Payment |
| **District of Columbia** | Coverage | Coverage | Coverage | Coverage |
| **Florida** | None | None | None | None |
| **Georgia** | None | None | Both | Both |
| **Hawaii** | Payment | Payment | Payment | Payment |
| **Idaho** | None | None | None | None |
| **Illinois** | None | None | None | Both |
| **Indiana** | Coverage | Coverage | Coverage | Coverage |
| **Iowa** | None | Coverage | Coverage | Coverage |
| **Kansas** | None | None | None | None |
| **Kentucky** | None | Both | Both | Both |
| **Louisiana** | None | None | None | Coverage |
| **Maine** | Coverage | Coverage | Coverage | Coverage |
| **Maryland** | None | None | Coverage | Both |
| **Massachusetts** | Coverage | Coverage | Coverage | Coverage |
| **Michigan** | None | None | None | None |
| **Minnesota** | Both | Both | Both | Coverage |
| **Mississippi** | Coverage | Coverage | Coverage | Both |
| **Missouri** | Both | Both | Both | Both |
| **Montana** | Coverage | Coverage | Coverage | Coverage |
| **Nebraska** | Coverage | Coverage | Coverage | Coverage |
| **Nevada** | Coverage | Coverage | Coverage | Both |
| **New Hampshire** | Coverage | Coverage | Both | Both |
| **New Jersey** | Both | Both | Both | Both |
| **New Mexico** | Coverage | Both | Both | Coverage |
| **New York** | Coverage | Coverage | Coverage | Coverage |
| **North Carolina** | None | None | None | None |
| **North Dakota** | Coverage | Coverage | Coverage | Coverage |
| **Ohio** | None | Coverage | Coverage | Coverage |
| **Oklahoma** | None | None | None | Both |
| **Oregon** | Coverage | Coverage | Coverage | Coverage |
| **Pennsylvania** | None | None | None | None |
| **Rhode Island** | Coverage | Coverage | Both | Both |
| **South Carolina** | None | None | None | None |
| **South Dakota** | None | Coverage | Coverage | Coverage |
| **Tennessee** | Coverage | Coverage | Coverage | Coverage |
| **Texas** | None | None | None | None |
| **Utah** | None | None | None | Coverage |
| **Vermont** | Coverage | Coverage | Coverage | Coverage |
| **Virginia** | Both | Both | Both | Both |
| **Washington** | None | None | Payment | Payment |
| **West Virginia** | None | None | Coverage | Coverage |
| **Wisconsin** | None | None | None | None |
| **Wyoming** | None | None | None | None |

Note: *None* indicates no relevant telehealth payment or coverage parity laws identified during the respective years. *Both* indicates telehealth payment and coverage parity laws were in effect during the respective years.

**Appendix Table 4.** Identification of outpatient telehealth encounters.

| **Description** | **Value** |
| --- | --- |
| **Place of Service** | =2<br>(i.e., telehealth) |
| **Procedure Modifier** |  |
| **Synchronous telemedicine service rendered via telephone or other real-time interactive audio-only telecommunications system** | =93 |
| **Synchronous telemedicine service rendered via real-time interactive audio and visual telecommunication system** | =95 |
| **Telehealth services for diagnosis, evaluation, or treatment of symptoms of acute stroke** | =GO |
| **Telehealth service rendered via asynchronous telecommunications system** | =GQ |
| **Telehealth service rendered via interactive audio and video telecommunication systems** | =GT |

**Appendix Table 5.** Distribution of sample by 50 states and the District of Columbia.

| **States** | **Freq.** | **Percent %** |
| --- | --- | --- |
| **Alabama** | 23,499 | 6.65 |
| **Alaska** | 169 | 0.05 |
| **Arizona** | 6,659 | 1.89 |
| **Arkansas** | 1,663 | 0.47 |
| **California** | 12,967 | 3.67 |
| **Colorado** | 2,552 | 0.72 |
| **Connecticut** | 3,577 | 1.01 |
| **Delaware** | 1,680 | 0.48 |
| **Florida** | 22,778 | 6.45 |
| **Georgia** | 22,904 | 6.48 |
| **Hawaii** | 38 | 0.01 |
| **Idaho** | 381 | 0.11 |
| **Illinois** | 11,347 | 3.21 |
| **Indiana** | 10,754 | 3.04 |
| **Iowa** | 1,313 | 0.37 |
| **Kansas** | 2,449 | 0.69 |
| **Kentucky** | 22,911 | 6.49 |
| **Louisiana** | 3,577 | 1.01 |
| **Maine** | 1,100 | 0.31 |
| **Maryland** | 6,254 | 1.77 |
| **Massachusetts** | 7,049 | 2.00 |
| **Michigan** | 24,050 | 6.81 |
| **Minnesota** | 1,865 | 0.53 |
| **Mississippi** | 5,024 | 1.42 |
| **Missouri** | 6,532 | 1.85 |
| **Montana** | 296 | 0.08 |
| **Nebraska** | 709 | 0.20 |
| **Nevada** | 903 | 0.26 |
| **New Hampshire** | 740 | 0.21 |
| **New Jersey** | 9,713 | 2.75 |
| **New Mexico** | 328 | 0.09 |
| **New York** | 12,160 | 3.44 |
| **North Carolina** | 14,250 | 4.03 |
| **North Dakota** | 174 | 0.05 |
| **Ohio** | 21,958 | 6.22 |
| **Oklahoma** | 2,739 | 0.78 |
| **Oregon** | 5,117 | 1.45 |
| **Pennsylvania** | 13,663 | 3.87 |
| **Rhode Island** | 515 | 0.15 |
| **South Carolina** | 4,741 | 1.34 |
| **South Dakota** | 572 | 0.16 |
| **Tennessee** | 9,541 | 2.70 |
| **Texas** | 29,030 | 8.22 |
| **Utah** | 800 | 0.23 |
| **Vermont** | 478 | 0.14 |
| **Virginia** | 12,818 | 3.63 |
| **Washington** | 2,975 | 0.84 |
| **District of Columbia** | 441 | 0.12 |
| **West Virginia** | 2,140 | 0.61 |
| **Wisconsin** | 3,119 | 0.88 |
| **Wyoming** | 208 | 0.06 |
| **Total** | 353,220 | 100 |

**Appendix Table 6.** The association of <u>telehealth payment parity</u> laws with number of hypertension-related telehealth and in-person visits.^a^.

|  | **Number of**<br>**hypertension-related**<br>**telehealth visits per**<br>**1,000 patients** | **Number of CVD-**<br>**related telehealth**<br>**visits per 1,000**<br>**patients** |
| --- | --- | --- |
| Payment parity | 2.61**<br>(0.99 - 4.23) | 0.92**<br>(0.23 - 1.61) |
| Covariates |  |  |
| COVID-19 diagnosis | 111.80***<br>(107.80 - 115.80) | 21.13***<br>(19.36 - 22.90) |
| Number of in-person visits | 0.48***<br>(0.44 - 0.52) | 0.29***<br>(0.27 - 0.31) |
| Aged 18–34 [reference] |  |  |
| Aged 35–44 | 5.89**<br>(2.35 - 9.43) | 7.63***<br>(5.48 - 9.79) |
| Aged 45–54 | -3.90*<br>(-7.25 - -0.55) | 13.32***<br>(11.14 - 15.50) |
| Aged 55–64 | -8.93***<br>(-12.38 - -5.48) | 18.62***<br>(16.26 - 20.97) |
| Male [reference] |  |  |
| Female | 14.79***<br>(13.41 - 16.16) | -5.55***<br>(-6.24 - -4.85) |
| Rural [reference] |  |  |
| Urban | 31.22***<br>(29.08 - 33.36) | 6.51***<br>(5.53 - 7.49) |
| Comorbidities |  |  |
| Myocardial infarction | -7.53**<br>(-12.72 - -2.34) | 22.05***<br>(20.10 - 24.00) |
| Congestive heart failure | 2.76<br>(-1.51 - 7.04) | 26.87***<br>(24.77 - 28.97) |
| Cerebrovascular | 5.84*<br>(1.17 - 10.51) | 18.36***<br>(16.60 - 20.13) |
| Peripheral vascular | 6.67<br>(-0.87 - 14.21) | 16.22***<br>(13.79 - 18.65) |
| Dementia | -4.72<br>(-42.64 - 33.20) | 3.72<br>(-9.24 - 16.68) |
| Chronic pulmonary disease | 12.18***<br>(9.82 - 14.53) | 5.98***<br>(4.98 - 6.97) |
| Rheumatic disease | 15.21***<br>(10.44 - 19.99) | 10.97***<br>(9.10 - 12.84) |
| Peptic ulcer disease | -3.82<br>(-14.43 - 6.78) | -2.04<br>(-5.99 - 1.92) |
| Mild liver disease | 6.08** | 0.13 |
|  | (2.21 - 9.94) | (-1.45 - 1.72) |
| Diabetes without chronic complication | 18.25*** | 1.77*** |
|  | (16.54 - 19.95) | (1.05 - 2.49) |
| Diabetes with chronic complication | 11.84*** | 3.55*** |
|  | (8.72 - 14.95) | (2.33 - 4.78) |
| Hemiplegia or paraplegia | 3.80 | 10.35*** |
|  | (-7.92 - 15.51) | (6.94 - 13.76) |
| Renal disease | 57.78*** | 0.30 |
|  | (54.15 - 61.40) | (-1.05 - 1.64) |
| Any malignancy | 0.97 | 1.50* |
|  | (-2.70 - 4.63) | (0.03 - 2.97) |
| Moderate or severe liver disease | -12.95 | 0.43 |
|  | (-30.21 - 4.30) | (-5.34 - 6.19) |
| Metastatic solid tumor | 5.38 | -0.92 |
|  | (-6.27 - 17.02) | (-5.44 - 3.61) |
| AIDS/HIV | 64.51*** | 7.53*** |
|  | (54.48 - 74.53) | (3.18 - 11.88) |
| State-year level covariates |  |  |
| Unemployment rates | -0.13 | -0.12 |
|  | (-0.94 - 0.67) | (-0.45 - 0.22) |
| GDP per capita | -0.00*** | -0.00 |
|  | (-0.00 - -0.00) | (-0.00 - 0.00) |
| Poverty rates | 0.24 | 0.09 |
|  | (-0.22 - 0.70) | (-0.11 - 0.29) |
| State fixed effects | Yes | Yes |
| Year fixed effects | Yes | Yes |
| Observations | 353,220 | 353,220 |

Abbreviation: CVD, cardiovascular disease.

**Appendix Table 7.** The association of telehealth <u>coverage parity laws</u> with number of hypertension-related telehealth and in-person visits.^a^.

|  | **Number of**<br>**hypertension-related**<br>**telehealth visits per**<br>**1,000 patients** | **Number of CVD-**<br>**related telehealth**<br>**visits per 1,000**<br>**patients** |
| --- | --- | --- |
| Coverage parity | 1.49<br>(-0.47 - 3.45) | 0.76<br>(-0.04 - 1.56) |
| Covariates |  |  |
| COVID-19 diagnosis | 111.80***<br>(107.80 - 115.70) | 21.09***<br>(19.33 - 22.85) |
| Number of in-person visits | 0.48***<br>(0.44 - 0.52) | 0.29***<br>(0.27 - 0.31) |
| Aged 18–4 [reference] |  |  |
| Aged 35–44 | 5.89**<br>(2.35 - 9.43) | 7.62***<br>(5.47 - 9.77) |
| Aged 45–54 | -3.90*<br>(-7.25 - -0.54) | 13.30***<br>(11.13 - 15.47) |
| Aged 55–64 | -8.93***<br>(-12.38 - -5.47) | 18.58***<br>(16.24 - 20.93) |
| Male [reference] |  |  |
| Female | 14.79***<br>(13.41 - 16.16) | -5.54***<br>(-6.23 - -4.84) |
| Rural [reference] |  |  |
| Urban | 31.22***<br>(29.08 - 33.36) | 6.49***<br>(5.52 - 7.47) |
| Comorbidities |  |  |
| Myocardial infarction | -7.53**<br>(-12.72 - -2.34) | 22.01***<br>(20.06 - 23.95) |
| Congestive heart failure | 2.77<br>(-1.50 - 7.04) | 26.83***<br>(24.74 - 28.92) |
| Cerebrovascular | 5.83*<br>(1.16 - 10.49) | 18.33***<br>(16.57 - 20.08) |
| Peripheral vascular | 6.68<br>(-0.86 - 14.22) | 16.19***<br>(13.76 - 18.61) |
| Dementia | -4.71<br>(-42.63 - 33.21) | 3.70<br>(-9.24 - 16.63) |
| Chronic pulmonary disease | 12.17***<br>(9.82 - 14.53) | 5.97***<br>(4.98 - 6.96) |
| Rheumatic disease | 15.22***<br>(10.44 - 19.99) | 10.95***<br>(9.08 - 12.82) |
| Peptic ulcer disease | -3.82<br>(-14.42 - 6.78) | -2.04<br>(-5.99 - 1.91) |
| Mild liver disease | 6.08** | 0.14 |
|  | (2.22 - 9.94) | (-1.45 - 1.72) |
| Diabetes without chronic complication | 18.25*** | 1.77*** |
|  | (16.54 - 19.95) | (1.05 - 2.49) |
| Diabetes with chronic complication | 11.83*** | 3.54*** |
|  | (8.72 - 14.95) | (2.32 - 4.77) |
| Hemiplegia or paraplegia | 3.78 | 10.33*** |
|  | (-7.93 - 15.49) | (6.92 - 13.73) |
| Renal disease | 57.78*** | 0.30 |
|  | (54.15 - 61.40) | (-1.05 - 1.64) |
| Any malignancy | 0.97 | 1.50* |
|  | (-2.70 - 4.64) | (0.03 - 2.97) |
| Moderate or severe liver disease | -12.97 | 0.43 |
|  | (-30.22 - 4.29) | (-5.32 - 6.18) |
| Metastatic solid tumor | 5.38 | -0.92 |
|  | (-6.27 - 17.02) | (-5.43 - 3.60) |
| AIDS/HIV | 64.51*** | 7.53*** |
|  | (54.49 - 74.54) | (3.19 - 11.86) |
| State-year level covariates |  |  |
| Unemployment rates | 0.08 | -0.04 |
|  | (-0.72 - 0.88) | (-0.37 - 0.29) |
| GDP per capita | -0.00** | -0.00 |
|  | (-0.00 - -0.00) | (-0.00 - 0.00) |
| Poverty rates | 0.26 | 0.09 |
|  | (-0.21 - 0.74) | (-0.12 - 0.29) |
| State fixed effects | Yes | Yes |
| Year fixed effects | Yes | Yes |
| Observations | 353,220 | 353,220 |

Abbreviation: CVD, cardiovascular disease.

**Appendix Table 8.** Sensitivity analyses of the association of telehealth payment and coverage parity laws with medication adherence, 2018–2021.^a^.

| Sensitivity analysis | Probit | OLS | Probit | OLS |
| --- | --- | --- | --- | --- |
|  | Medication Adherence | Medication Adherence | Medication Adherence | Medication Adherence |
| Payment parity | **0.46\***<br>**(0.01- 0.92)** | 0.46*<br>(0.01 - 0.92) | **NA** | **NA** |
| Coverage parity | **NA** |  | 0.10<br>(-0.37 - 0.57) | 0.10<br>(-0.37 - 0.57) |
| Covariates |  |  |  |  |
| COVID-19 diagnosis | -3.60***<br>(-4.06 - -3.15) | -3.60***<br>(-4.06 - -3.15) | -3.61***<br>(-4.06 - -3.15) | -3.61***<br>(-4.06 - -3.15) |
| Number of in-person visits | -0.00<br>(-0.02 - 0.02) | -0.00<br>(-0.02 - 0.02) | -0.00<br>(-0.02 - 0.02) | -0.00<br>(-0.02 - 0.02) |
| Aged 18–34 [reference] |  |  |  |  |
| Aged 35–44 | 9.69***<br>(8.84 – 10.50) | 9.69***<br>(8.84 - 10.53) | 9.69***<br>(8.84 – 10.50) | 9.69***<br>(8.84 - 10.53) |
| Aged 45–54 | 18.50***<br>(17.70 – 19.30) | 18.53***<br>(17.74 - 19.33) | 18.50***<br>(17.70 – 19.30) | 18.53***<br>(17.74 - 19.33) |
| Aged 55–64 | 24.40***<br>(23.50 – 25.40) | 24.44***<br>(23.48 - 25.39) | 24.40***<br>(23.50 – 25.40) | 24.44***<br>(23.48 - 25.39) |
| Male [reference] |  |  |  |  |
| Female | -3.96***<br>(-4.41 - -3.51) | -3.96***<br>(-4.41 - -3.51) | -3.96***<br>(-4.41 - -3.51) | -3.96***<br>(-4.41 - -3.51) |
| Rural [reference] |  |  |  |  |
| Urban | -0.41<br>(-1.10 - 0.28) | -0.41<br>(-1.10 - 0.28) | -0.41<br>(-1.10 - 0.28) | -0.41<br>(-1.10 - 0.28) |
| Comorbidities |  |  |  |  |
| Myocardial infarction | -0.81<br>(-1.94 - 0.33) | -0.81<br>(-1.94 - 0.33) | -0.81<br>(-1.94 - 0.33) | -0.81<br>(-1.94 - 0.33) |
| Congestive heart failure | -2.90***<br>(-3.77 - -2.03) | -2.90***<br>(-3.77 - -2.03) | -2.90***<br>(-3.77 - -2.03) | -2.90***<br>(-3.77 - -2.03) |
| Cerebrovascular | -0.85 | -0.85 | -0.85 | -0.85 |
|  | (-1.75 - 0.04) | (-1.75 - 0.04) | (-1.75 - 0.04) | (-1.75 - 0.04) |
| Peripheral vascular | -3.01*** | -3.01*** | -3.01*** | -3.01*** |
|  | (-4.56 - -1.46) | (-4.56 - -1.46) | (-4.56 - -1.46) | (-4.56 - -1.46) |
| Dementia | -10.20** | -10.22** | -10.2** | -10.22** |
|  | (-17.30 - -3.15) | (-17.29 - -3.15) | (-17.30 - -3.15) | (-17.29 - -3.15) |
| Chronic pulmonary disease | -3.50*** | -3.50*** | -3.50*** | -3.50*** |
|  | (-4.05 - -2.95) | (-4.05 - -2.95) | (-4.05 - -2.95) | (-4.05 - -2.95) |
| Rheumatic disease | -2.30*** | -2.30*** | -2.30*** | -2.30*** |
|  | (-3.39 - -1.22) | (-3.39 - -1.22) | (-3.39 - -1.22) | (-3.39 - -1.22) |
| Peptic ulcer disease | -9.59*** | -9.59*** | -9.59*** | -9.59*** |
|  | (-11.0 - -7.84) | (-11.34 - -7.84) | (-11.3 - -7.84) | (-11.34 - -7.84) |
| Mild liver disease | -2.44*** | -2.44*** | -0.0244*** | -2.44*** |
|  | (-3.41 - -1.47) | (-3.41 - -1.47) | (-3.41 - -1.47) | (-3.41 - -1.47) |
| Diabetes without chronic complication | 1.45*** | 1.45*** | 1.45*** | 1.45*** |
|  | (1.05 - 1.84) | (1.05 - 1.84) | (1.05 - 1.84) | (1.05 - 1.84) |
| Diabetes with chronic complication | -2.04*** | -2.04*** | -2.03*** | -2.03*** |
|  | (-2.68 - -1.39) | (-2.68 - -1.39) | (-2.68 - -1.39) | (-2.68 - -1.39) |
| Hemiplegia or paraplegia | -2.78* | -2.78* | -2.78* | -2.78* |
|  | (-5.35 - -2.21) | (-5.35 - -0.22) | (-5.35 - -0.22) | (-5.35 - -0.22) |
| Renal disease | -2.02*** | -2.02*** | -2.02*** | -2.02*** |
|  | (-2.75 - -1.29) | (-2.75 - -1.29) | (-2.75 - -1.29) | (-2.75 - -1.29) |
| Any malignancy | 1.63*** | 1.63*** | 1.63*** | 1.63*** |
|  | (0.82 - 2.45) | (0.82 - 2.45) | (0.82 - 2.45) | (0.82 - 2.45) |
| Moderate or severe liver disease | -10.2*** | -10.21*** | -10.2*** | -10.21*** |
|  | (-14.40 - -6.04) | (-14.37 - -6.04) | (-14.40 - -6.04) | (-14.37 - -6.04) |
| Metastatic solid tumor | -7.18*** | -7.18*** | -7.18*** | -7.18*** |
|  | (-9.14 - -5.22) | (-9.14 - -5.22) | (-9.14 - -5.22) | (-9.14 - -5.22) |
| AIDS/HIV | 2.94** | 2.94** | 2.94** | 2.94** |
|  | (1.16 - 4.72) | (1.16 - 4.72) | (1.16 - 4.72) | (1.16 - 4.72) |
| State-year level covariates |  |  |  |  |
| Unemployment rates | 0.31* | 0.31* | 0.30* | 0.29* |
|  | (0.06 - 0.56) | (0.06 - 0.56) | (0.04 - 0.55) | (0.04 - 0.55) |
| GDP per capita | -0.00 | -0.00 | -0.00 | -0.00 |
|  | (-0.00 - 0.00) | (-0.00 - 0.00) | (-0.00 - 0.00) | (-0.00 - 0.00) |
| Poverty rates | 0.04 | 0.04 | 0.03 | 0.03 |
|  | (-0.11 - 0.19) | (-0.11 - 0.19) | (-0.12 - 0.18) | (-0.12 - 0.18) |
| State fixed effects | Yes | Yes | Yes | Yes |
| Year fixed effects | Yes | Yes | Yes | Yes |
| Observations | 353,220 | 353,220 | 353,220 | 353,220 |

Abbreviation: NA, not available.
<sup>a</sup> A probit and OLS were used as sensitivity analyses for the medication adherence dependent variable. All models were adjusted for state-fixed effects, year-fixed effects, COVID-19 diagnosis, number of in-person visits, patients' age groups, sex, urbanicity of residence, comorbidities, and state-year level time-varying covariates (state-year level unemployment rates, GDP per capita, and poverty rates). Average marginal effects with 95% CI (in parentheses) were reported. All standard errors were clustered by states.

**Appendix Table 9.** Sensitivity analyses of the association of telehealth payment and coverage parity laws with average number of drug supply per antihypertensive drug, 2018–2021.^a^.

|  | **ZINB** | **ZINB** |
| --- | --- | --- |
|  | Average days of drug supply | Average days of drug supply |
| **Payment parity** | 1.88 | **NA** |
|  | (-0.11 - 3.86) |  |
| **Coverage parity** | **NA** | 1.92 |
|  |  | (-0.01 - 3.86) |
| **Covariates** |  |  |
| **COVID-19 diagnosis** | -3.49*** | -3.49*** |
|  | (-4.47 - -2.51) | (-4.48 - -2.51) |
| **Number of in-person visits** | -0.10*** | -0.10*** |
|  | (-0.13 - -0.07) | (-0.13 - -0.07) |
| **Aged 18–34 [reference]** |  |  |
| **Aged 35–44** | 12.47*** | 12.47*** |
|  | (11.38 - 13.56) | (11.38 - 13.56) |
| **Aged 45–54** | 21.95*** | 21.95*** |
|  | (20.51 - 23.39) | (20.51 - 23.39) |
| **Aged 55–64** | 26.74*** | 26.74*** |
|  | (24.68 - 28.79) | (24.68 - 28.80) |
| **Male [reference]** |  |  |
| **Female** | -9.14*** | -9.14*** |
|  | (-10.10 - -8.19) | (-10.10 - -8.19) |
| **Rural [reference]** |  |  |
| **Urban** | 6.23*** | 6.23*** |
|  | (3.01 - 9.45) | (3.01 - 9.45) |
| **Comorbidities** |  |  |
| **Myocardial infarction** | -18.62*** | -18.62*** |
|  | (-20.16 - -17.08) | (-20.16 - -17.08) |
| **Congestive heart failure** | -23.53*** | -23.53*** |
|  | (-25.22 - -21.84) | (-25.22 - -21.84) |
| **Cerebrovascular** | -10.40*** | -10.39*** |
|  | (-11.56 - -9.231) | (-11.56 - -9.23) |
| **Peripheral vascular** | -12.99*** | -12.99*** |
|  | (-15.30 - -10.67) | (-15.30 - -10.67) |
| **Dementia** | -29.33*** | -29.33*** |
|  | (-39.33 - -19.33) | (-39.33 - -19.34) |
| **Chronic pulmonary disease** | -6.49*** | -6.49*** |
|  | (-7.39 - -5.58) | (-7.39 - -5.58) |
| **Rheumatic disease** | -4.91*** | -4.91*** |
|  | (-6.74 - -3.09) | (-6.74 - -3.09) |
| **Peptic ulcer disease** | -15.25*** | -15.24*** |
|  | (-18.37 - -12.12) | (-18.37 - -12.11) |
| **Mild liver disease** | -5.91*** | -5.91*** |
|  | (-7.60 - -4.23) | (-7.60 - -4.23) |
| **Diabetes without chronic complication** | 8.54*** | 8.54*** |
|  | (7.76 - 9.33) | (7.76 - 9.33) |
| **Diabetes with chronic complication** | -1.71*** | -1.71*** |
|  | (-2.46 - -0.96) | (-2.46 - -0.96) |
| **Hemiplegia or paraplegia** | -12.89*** | -12.88*** |
|  | (-18.18 - -7.60) | (-18.18 - -7.59) |
| **Renal disease** | -13.87*** | -13.86*** |
|  | (-15.77 - -11.96) | (-15.77 - -11.96) |
| **Any malignancy** | -0.28 | -0.28 |
|  | (-1.65 - 1.10) | (-1.65 - 1.10) |
| **Moderate or severe liver disease** | -35.11*** | -35.11*** |
|  | (-40.82 - -29.41) | (-40.82 - -29.41) |
| **Metastatic solid tumor** | -12.35*** | -12.35*** |
|  | (-16.96 - -7.74) | (-16.96 - -7.74) |
| **AIDS/HIV** | -8.00*** | -8.00*** |
|  | (-11.93 - -4.07) | (-11.93 - -4.07) |
| **State-year level covariates** |  |  |
| **Unemployment rates** | -0.28 | -0.22 |
|  | (-1.00 - 0.43) | (-0.92 - 0.49) |
| **GDP per capita** | -0.00 | -0.00 |
|  | (-0.00 - 0.00) | (-0.00 - 0.00) |
| **Poverty rates** | -0.21 | -0.29* |
|  | (-0.44 - 0.02) | (-0.51 - -0.06) |
| **State fixed effects** | Yes | Yes |
| **Year fixed effects** | Yes | Yes |
| **Observations** | 353,220 | 353,220 |

Abbreviations: ZINB, zero-inflated negative binomial; NA, not available.
<sup>a</sup> A zero-inflated negative binomial model was used for sensitivity analysis. The logit model was used to model zero outcomes (first part). A negative binomial model was used to model the number of average days of drug supply (second part). The first part was adjusted for COVID-19 diagnosis, number of in-person visits, patients' age groups, sex, urbanicity of residence, and comorbidities. The second part was adjusted for state-fixed effects, year-fixed effects, COVID-19 diagnosis, number of in-person visits, patients' age groups, sex, urbanicity of residence, comorbidities, and state-year level time-varying covariates (state-year level unemployment rates, GDP per capita, and poverty rates). Average marginal effects with 95% CI were reported. All standard errors were clustered by states.

